# Multidimensional assessment of adverse events of finasteride: a real-world pharmacovigilance analysis based on FDA Adverse Event Reporting System (FAERS) from 2004 to April 2024

**DOI:** 10.1101/2024.08.21.24312383

**Authors:** Xiaoling Zhong, Yihan Yang, Sheng Wei, Yuchen Liu

**Affiliations:** Guangdong Provincial Hospital of Traditional Chinese Medicine, Guangzhou, China; The Institution of Rehabilitation Industry, Fujian University of Traditional Chinese Medicine, Fuzhou, China; The Second Affiliated Hospital of Wannan Medical College, Wuhu, China; Shenzhen Luohu District Hospital of Traditional Chinese Medicine, Shenzhen, China

**Author notes:** Corresponding author (YL). These authors contributed equally to this work (Xiaoling Zhong, Yihan Yang).

**Keywords:** real-world pharmacovigilance analysis, FAERS, pharmacovigilance, finasteride

## Abstract

**Background:** Finasteride is commonly utilized in clinical practice for treating androgenetic alopecia, but real-world data regarding the long-term safety of its adverse events remains incomplete, necessitating ongoing supplementation. This study aims to evaluate the adverse events (AEs) associated with finasteride use, based on data from the US Food and Drug Administration Adverse Event Reporting System (FAERS), to contribute to its safety assessment.

**Methods:** We reviewed adverse event reports associated with finasteride from the FAERS database, covering the period from the first quarter of 2004 to the first quarter of 2024. We assessed the safety of finasteride medication and AEs using four proportional disproportionality analyses: reported odds ratio, proportionate reporting ratio (PRR), Bayesian Confidence Propagation Neural Network (BCPN), and Multi-Item Gamma Poisson Shrinkage (MGPS). These methods were used to evaluate the of finasteride medication and AEs. whether there is a significant association between finasteride drug use and AEs. To investigate potential safety issues related to drug use, we further analyzed the similarities and differences in the onset time and AEs by gender, as well as the similarities and differences in AEs by age.

**Results:** Among the 11,557 adverse event reports where finasteride was the primary suspected drug, most patients affected were male (86.04%), with a significant proportion being the young adult aged 18-45 years (27.22%). We categorized 73 adverse events (AEs) into 7 different system organ categories (SOCs), which included common AEs like erectile dysfunction and sexual dysfunction. Notably, Peyronie’s disease and post 5α reductase inhibitor syndrome were AEs not listed in the drug insert. We identified 102 AEs for men and 7 for women. Depression and anxiety were notable AEs for both male and female. Additionally, we examined 17 adverse events (AEs) in patients under 18 years old, 157 in patients aged 18 to 65 years, and 133 in patients aged 65 years and older. Each age group exhibited unique AEs, although erectile dysfunction, decreased libido, depression, suicidal ideation, psychotic disorder, and attention disturbance were common AEs observed across different age brackets. Ultimately, the median onset time for all instances was 61 days. The onset was mainly within one month after initiation of finasteride and it is noteworthy that the second highest number of cases involved adverse drug reactions persisted beyond one year of treatment.

**Conclusion:** The results of our study uncovered both known and novel AEs associated with finasteride medication. Some of these AEs were identical to the specification, and some of them signaled AEs that were not demonstrated in the specification. In addition, some AEs showed variations based on gender and age in our study. Consequently, our findings offer valuable insights for future research on the safety of finasteride medication and are anticipated to enhance its safe use in clinical practice.

## Introduction

Androgenetic alopecia (AGA), commonly referred to as male-pattern baldness, is characterized by hair loss driven by dihydrotestosterone, a potent derivative of testosterone[1].It has a significant negative physiological and psychological impact on patients. Patients with AGA experience adverse effects such as sexual dysfunction, affective disorders, stigma and low self-esteem compared to normal individuals[2,3], especially in younger patients [4]. Fortunately, however, since receiving approval for treating AGA in 1997, finasteride has gained popularity for AGA treatment and has shown positive therapeutic outcomes[5].

Finasteride, as the main drug for the treatment of AGA, works by efficiently inhibiting 5-alpha reductase inhibitors[6]. Since the synthesis of sex hormones in the body is dependent on steroid reductases, and SRD5A2 is a biofilm chimeric steroid in the class of steroid reductase enzymes, SRD5A2 can synthesize dihydrotestosterone by catalyzing the reductive reaction of testosterone, which can lead to the onset of AGA if it is too high[7]. Therefore, it is important to note that when neuroactive steroids have a negative effect on mood, behavior and cognition, they are widely used as anti-androgens. So finasteride as 5-alpha reductase inhibitor is the anti-androgenic drug that is used to treat both diseases and is widely available[8]. It is noteworthy that patients with BPH and androgenetic alopecia experience adverse events of depression or suicide when neuroactive steroids have a reduced ability to control mood, behavior, and cognition[9–11]. The data show that between 1998 and 2008, an estimated 4.6 million men were treated with at least 1 mg of finasteride[12]. More importantly, as of June 2021, the FDA adverse event disclosure data have reported up to 10,295 serious adverse events with finasteride use. Unfortunately, however, safety information regarding finasteride in patients with androgenetic alopecia primarily stems from clinical trials and post-marketing observational studies. The frequency of persistent adverse events associated with it remains unclear, underscoring the necessity for additional evidence-based research to investigate the potential risks of finasteride use[13]. A Real-world study, with their large data samples and rigorous inclusion and exclusion criteria, have emerged as a useful tool to explore adverse events and to compensate for clinical trials. An effective way of exploring adverse drug events and bridging the gap between clinical trials to provide us with evidence-based information.

The U.S. Food and Drug Administration Adverse Event Reporting System (FAERS) is an openly available database created by the FDA that gathers instances of adverse drug events (AEs) reported globally, with a large amount of real-world data and extensive geographic coverage of records of AEs from practicing physicians, pharmacists, registered nurses, consumers, and other healthcare professionals, among others[14,15]. Being the largest pharmacovigilance database globally, FAERS is an invaluable resource for identifying adverse events associated with drug usage[16]. Thus, the objective of this study was to assess the long-term safety profile of finasteride following its market introduction using data extracted from the FDA Adverse Event Reporting System (FAERS) database[17]. This study offers guidance and citations for the rational and safe clinical administration of finasteride dosage.

## Materials and methods

### Date sources

FAERS is a database for public post-marketing safety monitoring of drugs, and this retrospective analysis of adverse drug reactions utilized data sourced from the FAERS database. Our study gathered data from the FAERS Quarterly Data Extract Files spanning from Q1 2004 (FDA approval of finasteride) through Q1 2024.The FAERS database is updated quarterly and is widely recognized internationally for its large volume of data and standardization, and all the data within the FAERS database can be freely downloaded (https://open.fda.gov/data/downloads/).

### Data extraction

The FAERS data file contains seven comprehensive datasets: drug information (DRUG), information on adverse events (REAC), information on patient demographics and management (DEMO), information on patient outcomes (OUTC), information on the source of the report (RPSR), date of initiation and termination of medication (THER), and information on the indication of the drug (INDI). Whereas, the role of the reporting drug in each case was indicated in the DRUG table through specific codes: PS (primary suspect drug), SS (secondary suspect drug), C (concomitant drug), and I (interacting drug)[18]. Major endpoints were identified as disability (DS), hospitalisation (HO), life-threatening (LT), death (DE), and need for intervention to prevent permanent injury/damage (RI), congenital anomalies (CA), and other serious medical events (OT). In this study, cases were identified by utilizing drug names listed in the DRUG file, and selections were determined based on PS outcomes.

Adverse events in the FAERS database were categorized using the international medical dictionary MedDRA, which organizes terms into a structural hierarchy comprising preferred term (PT), system organ class (SOC), high level group term (HLGT), high level term (HLT), and lowest level term (LLT).We meticulously identified individual adverse event (AE) reports linked to finasteride at the system organ class (SOC) and preferred term (PT) levels. Additionally, our study concentrated on analyzing the specific drugs documented in the drug files, namely finasteride (generic name) and proscar (trade name), with the aim of obtaining a standardization of the AEs associated with finasteride after a name change[19,20]. Finally, by means of the Medex UIMA 1.8.3 system the standardization of drug names[21,22]. Finally, by means of the Medex UIMA 1.8.3 system the standardization of drug names.

### Data cleaning

A retrospective pharmacovigilance investigation was undertaken to assess the post-market safety of finasteride. Data were sourced from the openly accessible FAERS database, encompassing raw FAERS data related to finasteride spanning from the first quarter of 2004 to the first quarter of 2024. All data retrieved from the US FDA website were imported into R (Version 4.2.2) for subsequent analysis. Duplicate reports were identified and eliminated in accordance with FDA guidance, applying this rigorous methodology aimed at avoiding duplicate entry of adverse event reports and ensuring further analysis following a thorough evaluation of the safety profile of finasteride[23]. The operation was performed in the following manner, selecting the PRIMARYID, CASEID, and FDA_DT fields of the DEMO table and sorting the data by CASEID, FDA_DT, and PRIMARYID; when reports with the same identical CASEID are available, the report with the largest FDA_DT value is retained; when records with the same CASEID and FDA_DT are available, the record with the largest PRIMARYID value is retained[24]. Not only that, but the erroneous case reports noted on the FDA website were deleted as suggested, and the final obtained clinical characteristics of the patients who were administered finasteride were obtained Clinical characteristics including gender, age, reporting region, reporter, time of reporting, and outcome were the relevant AE data results.

### Statistical analysis

The four-grid proportional imbalance method, known for its widespread application in pharmacovigilance studies, was employed for the disproportionality analysis investigating potential associations between finasteride and all reported AEs[25], as documented in S1 Table. Reported dominance ratio (ROR), proportional reporting ratio (PRR), information component (IC) in Bayesian confidence propagation neural network (BCPNN), and multinomial gamma poisson shrinker (MGPS) are the four main specific metrics computed using standard formulas for evaluating potential correlations involving finasteride and AEs[26].

In our study, the signals of the 4 AE recordings of the target drug were calculated, and a positive drug-associated AE signal was considered if it met at least one of the four algorithms (lower limit of 95% CI > 1, N ≥ 3; PRR ≥ 2, χ2 ≥ 4, N ≥ 3; IC025 > 0 or EBGM05 > 2) [27,28]. The methodologies and thresholds for the adverse event signals are detailed in S2 Table, while the procedural flowchart is depicted in Fig 1.

**Fig 1.**
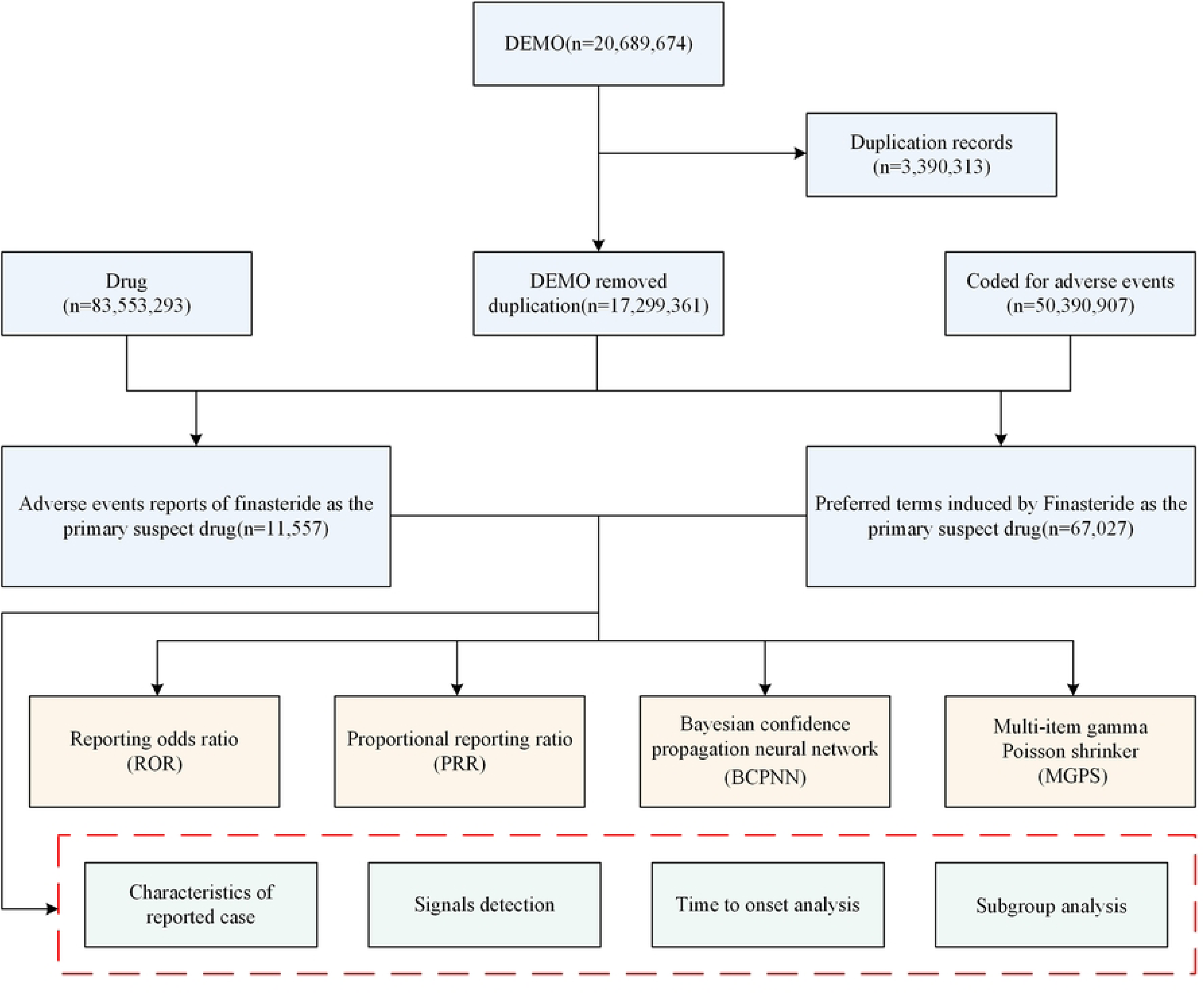
Flowchart depicting the selection of the study population.

The flow diagram of selecting finasteride therapy related AEs from the FAERS database. DEMO, demographic and administrative information; FAERS, US Food and Drug Administration Adverse Event Reporting System.

### Time to onset (TTO) analysis

The onset time of adverse events and the likelihood of a severe outcome were computed, with time to onset defined as the interval between EVENT_DT (date of AE onset) and START_DT (time of medication initiation), and median and interquartile ranges were used to describe the time to onset. In addition, AEs attributable to the route of administration were counted, calculated as number of serious outcomes/total number of events reported.

### Visualization of Data

Images were produced using the ggplot2 package and GraphPad Prism 8.0.1. We employed a global heat map to depict data from countries that submitted reports. A line graph was utilized to illustrate the trend in case numbers from the first quarter of 2004 to the first quarter of 2024. Additionally, to ascertain if the adverse event (AE) signal differed between males and females following finasteride administration, we created a volcano plot displaying log2-transformed PRP values on the x-axis and −log10-transformed adjusted P-values on the y-axis[24]. When PRP exceeded 1 and P.adj was greater than 0.05, it suggested a distinct AE signal between female and male patients. The demographic ratios of reported cases by sex and age, along with annual case counts, were processed and visualized using Excel tables.

### Ethics statement

The study involving human participants did not require ethical review and approval in compliance with local laws and institutional guidelines. Written informed consent from participants or their legally authorized representatives was not necessary for participation in this study in accordance with national regulations and institutional policies.

## Results

### Characteristics of reported case

From the first quarter of 2004 to the first quarter of 2024, there were 20,689,674 adverse reaction reports, of which 11,557 reports were adverse event reports involving the use of finasteride. Table 1 presents comprehensive details of the reports, encompassing patient demographics such as sex and age, the reporting year, the occupation of the reporter, and the country from which the report was submitted. The peak number of case reports for finasteride use was in 2024 (1141 9.87%), followed by 2015 (1000 8.65%) (Fig 2A). Geographical location information was not provided for 27.44% of all reporting countries (Fig 2B), which limited our insight into the relationship between geography and adverse events. The number of countries submitting reports included those from the United States (46.68%), followed by the United Kingdom (16.19%), Italy (3.51%), France (3.35%), and Germany (2.83%). Of the total reports (Fig 2C), 9944 (86.04 per cent) represented a larger proportion of males than females 397 (3.44 per cent), with the remaining 10.52 per cent being unknown. Moreover, Fig 2D reports are mainly submitted by consumers (53.67%) and physicians (20.73%). In terms of age (Fig 2E), the underlying audience for the drug was 18-45 years (27.22%), followed by 45-65 years (9.44%) and 7.81% of patients ≥75 years, with 48.80% of patients being of unknown age. After excluding product use for unknown indications and indications unknown, the most commonly reported indication was alopecia 3297 (28.36%), followed by androgenetic alopecia 1939 (16.68%) and benign prostatic hyperplasia 1053 (9.06%) (Fig 2F). Finally, the final outcome of the patients was disability in 22.60 per cent of cases and hospitalisation in 14.55 per cent of cases (Fig 2G).

**Fig 2.**
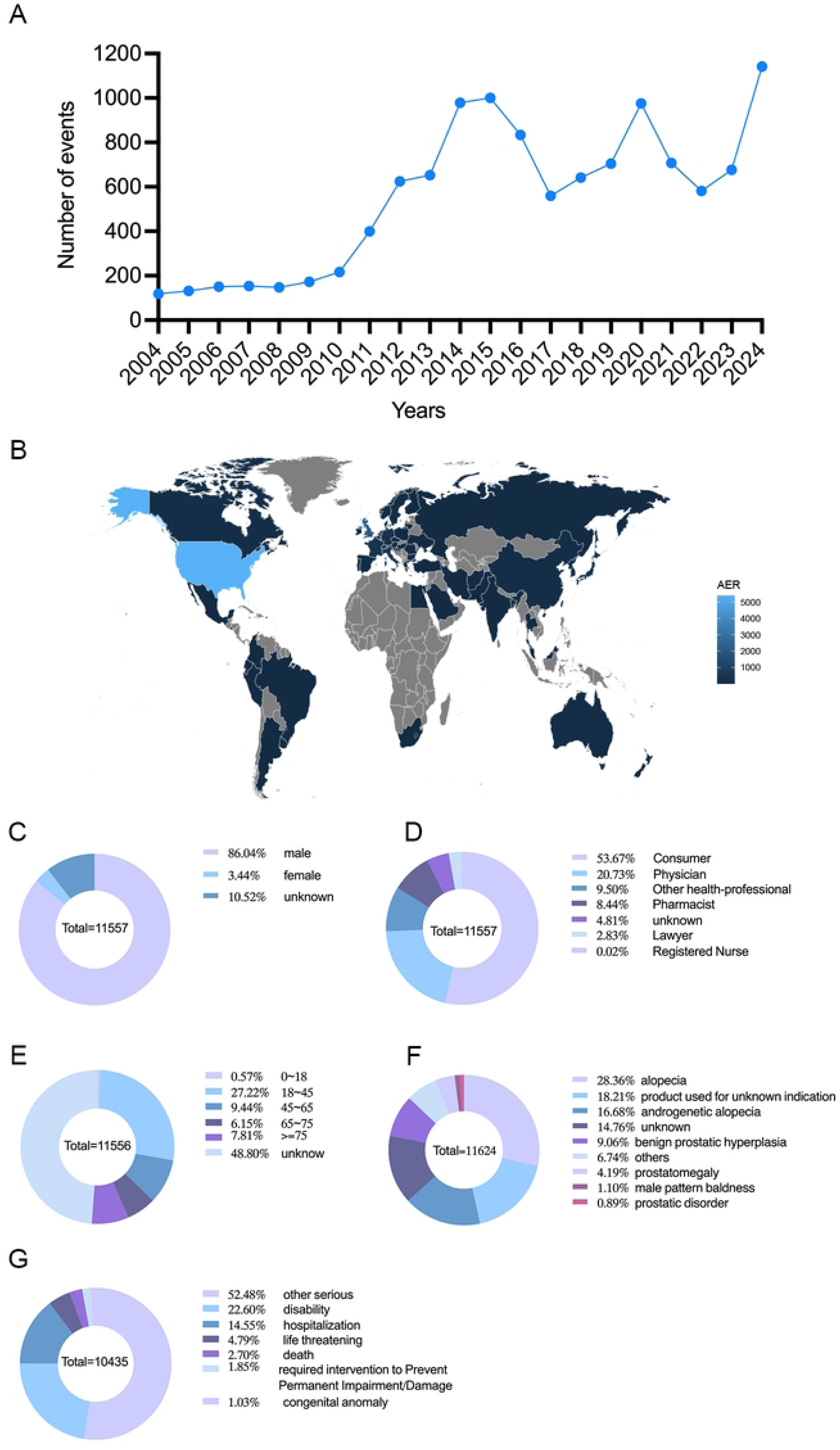
Basic information and patient characteristics according to the report. (A) The annual distribution of finasteride administration related AEs reports from 2004 to April 2024. (B) Country distribution of adverse events for finasteride administration, Darker colors represent a higher number of reports. (C) Gender ratio of male and female in reported events. (D) Occupational information ratio in reported events. (E) Age distribution ratio in reported events. (F) Ratio of indications in reported events. (G) Ratio of outcomes in reported events. Visualization through proportional area map, larger areas represent more reporters.

**Table 1.**
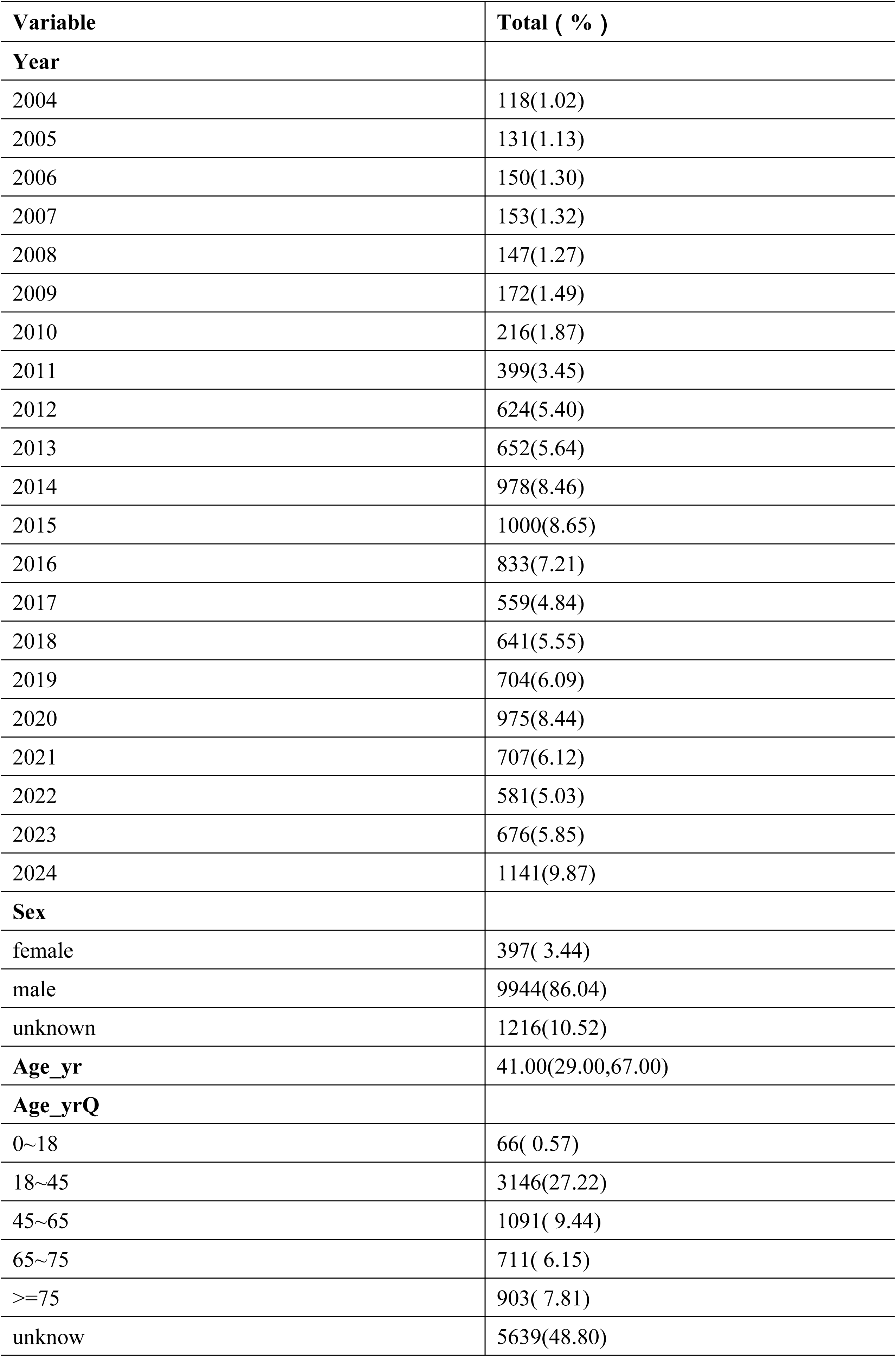
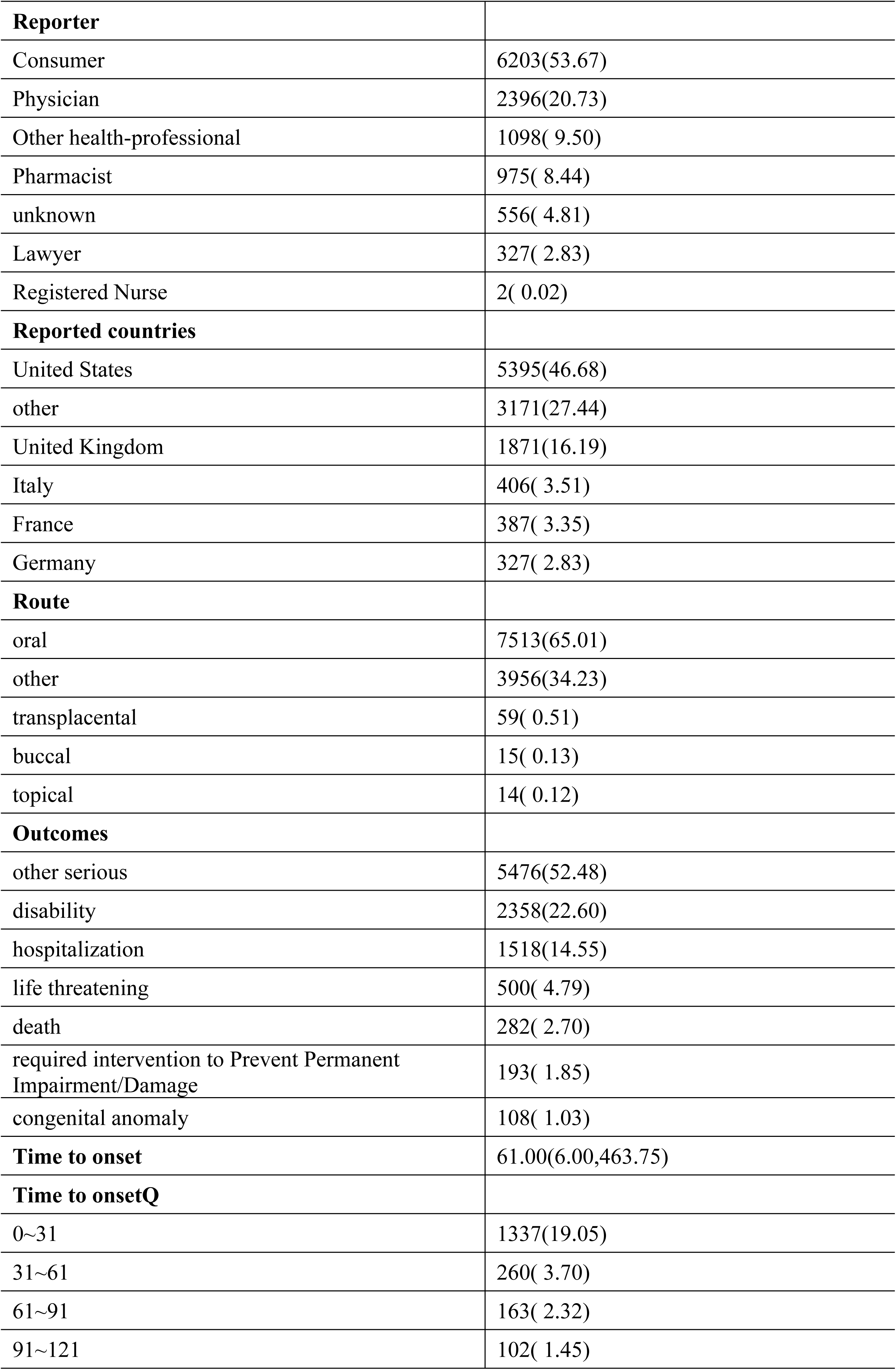
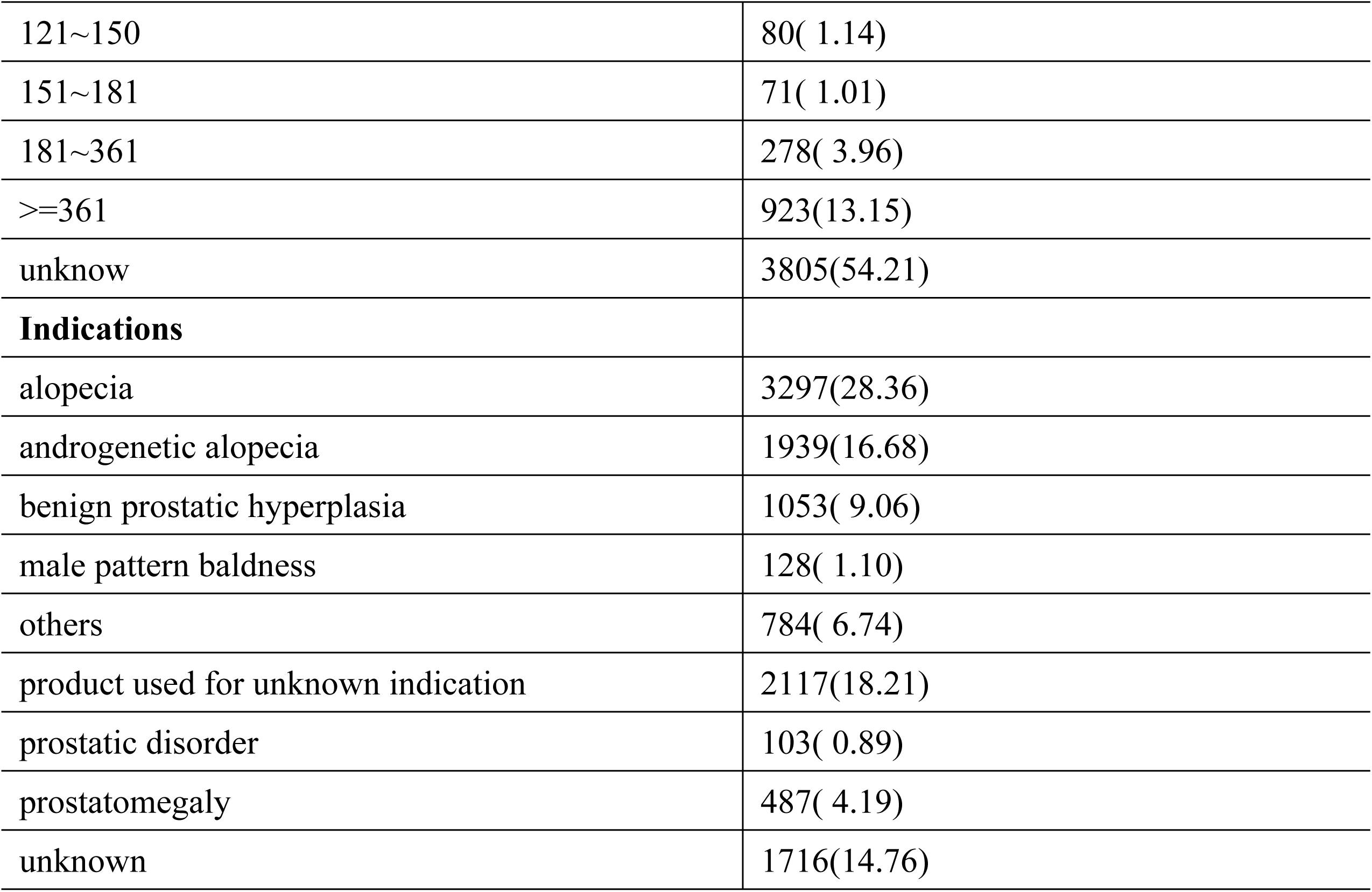
Clinical characteristics of reports with finasteride as the primary suspected drug from the FAERS database.

### Signals detection based on system organ class levels

Signals for finasteride at the SOC level are outlined in S3 Table, and our statistical evaluations indicated that finasteride dosing-related adverse events impacted a total of 24 SOCs. When we selected SOCs that met one of the four index criteria and ranked them in descending order by PRR ≥ 2, χ2 ≥ 4, and N ≥ 3, only reproductive system and breast disorders (ROR= 26.68, PRR= 21.97, χ2= 241136.36, IC= 4.42, EBGM= 21.37), endocrine disorders (ROR= 6.58, PRR= 6.49, χ2= 5179.48, IC= 2.69, EBGM= 6.44), psychiatric disorders (ROR= 4.57, PRR = 3.77, χ2= 32212.4, IC= 1.91, EBGM= 3.76) were eligible, indicating a high association. When we ranked the SOCs in descending order based on the number of cases, and the number of cases exceeded 200 were psychiatric disorders (n= 14951), reproductive and breast disorders (n= 12293), various neurological disorders (n= 5992), renal and urological disorders (n= 1841), various surgical and medical manipulations (n= 1268), and endocrine system disorders (n= 1122), various congenital familial genetic disorders (n= 282). These discoveries offer valuable understandings into particular domains where adverse events linked to finasteride are frequently documented.

We discovered that certain outcomes aligned with the SOC for common adverse reactions listed in the drug insert, demonstrating a high level of confidence in the data. The SOCs to which adverse reactions appeared in the insert included skin and subcutaneous tissue-like disorders (n= 2561, ROR= 0.68, PRR= 0.69, χ2= 369.34, IC= −0.53, EBGM= 0.69), neoplasms benign, malignant and unspecified (n= 1047, ROR= 0.56, PRR= 0.56, χ2= 364.75, IC= −0.83, EBGM= 0.56), but failed to demonstrate positive results at all four signal intensities, and their numerous reports may require further attention and research.

### Signals detection based on preferred term levels

In pharmacovigilance data analyses, we frequently utilize the PT terminology level to offer a comprehensive account of the precise clinical presentations, locations of occurrence, and specific disease types associated with an AE. We identified a total of 73 adverse events categorized at the PT-level based on one of the four screening methods outlined in Table 2.

**Table 2.** Signal strength of reports of finasteride administration at the preferred term level in the FAERS database.

| PTs and categories | Case Reports | ROR(95% CI) | PRR(95% CI) | chisq | IC(IC025) | EBGM(EBGM05) |
| --- | --- | --- | --- | --- | --- | --- |
| **reproductive system and breast disorders** |  |  |  |  |  |  |
| erectile dysfunction | 3377 | 144.15(138.85, 149.66) | 136.94(131.68, 142.41) | 385588.12 | 6.86(6.8) | 115.97(112.39) |
| sexual dysfunction | 2160 | 220.84(210.4, 231.79) | 213.75(205.53, 222.3) | 356099.85 | 7.38(7.31) | 166.6(159.99) |
| ejaculation disorder | 652 | 243.38(222.76, 265.92) | 241.02(222.85, 260.68) | 117980.64 | 7.51(7.39) | 182.7(169.65) |
| testicular pain | 618 | 223(203.8, 244) | 220.95(200.32, 243.7) | 104552.68 | 7.42(7.29) | 170.94(158.54) |
| ejaculation failure | 487 | 279.44(251.79, 310.14) | 277.42(251.52, 305.98) | 97945.39 | 7.66(7.52) | 202.84(185.9) |
| penile size reduced | 398 | 662.12(578.62, 757.67) | 658.19(573.81, 754.98) | 139167.42 | 8.46(8.28) | 351.19(313.73) |
| genital hypoaesthesia | 235 | 337.89(289.65, 394.18) | 336.71(287.84, 393.87) | 54305.2 | 7.86(7.65) | 232.77(204.62) |
| organic erectile dysfunction | 218 | 1094.72(889.08, 1347.92) | 1091.16(879.54, 1353.7) | 96782.38 | 8.8(8.56) | 445.36(374.2) |
| peyronie's disease | 215 | 286.11(244.49, 334.82) | 285.2(243.81, 333.62) | 44127.17 | 7.69(7.48) | 206.96(181.45) |
| testicular atrophy | 213 | 301.56(257.2, 353.56) | 300.6(256.98, 351.63) | 45419.19 | 7.75(7.53) | 214.94(188.15) |
| testicular disorder | 158 | 157.28(132.48, 186.73) | 156.91(131.53, 187.18) | 20246.36 | 7.02(6.78) | 129.96(112.58) |
| painful ejaculation | 96 | 707.65(535.46, 935.2) | 706.64(537.07, 929.76) | 34847.63 | 8.51(8.16) | 364.51(288.66) |
| varicocele | 89 | 461.45(354.37, 600.89) | 460.84(357.18, 594.58) | 25304.84 | 8.16(7.81) | 285.94(229.26) |
| male sexual dysfunction | 74 | 136.32(106.41, 174.65) | 136.17(105.54, 175.69) | 8405.1 | 6.85(6.5) | 115.42(93.81) |
| prostatic pain | 68 | 173.83(133.52, 226.32) | 173.65(134.59, 224.04) | 9480.26 | 7.14(6.77) | 141.22(113.25) |
| infertility male | 64 | 136.25(104.39, 177.84) | 136.12(103.46, 179.1) | 7266.87 | 6.85(6.47) | 115.38(92.33) |
| penile curvature | 63 | 328.78(244.5, 442.11) | 328.48(244.81, 440.75) | 14308.32 | 7.84(7.43) | 228.81(178.59) |
| male genital atrophy | 52 | 1116.34(727.21, 1713.69) | 1115.47(724.75, 1716.82) | 23293.43 | 8.81(8.32) | 449.35(313.93) |
| genital atrophy | 49 | 944(619.84, 1437.69) | 943.31(625.03, 1423.67) | 20441.53 | 8.71(8.21) | 418.62(294.41) |
| epididymal cyst | 49 | 800.35(535.17, 1196.92) | 799.77(529.92, 1207.04) | 18928.1 | 8.6(8.11) | 387.77(276.9) |
| genital paraesthesia | 49 | 230.1(167.07, 316.91) | 229.93(168.04, 314.62) | 8550.4 | 7.46(7.02) | 176.26(134.84) |
| scrotal disorder | 38 | 115.57(82.13, 162.64) | 115.51(82.78, 161.19) | 3738.49 | 6.65(6.16) | 100.24(75.32) |
| male reproductive tract disorder | 26 | 330.99(208.65, 525.06) | 330.86(206.71, 529.59) | 5935.04 | 7.85(7.22) | 229.96(156.31) |
| oligospermia | 23 | 239.92(150.02, 383.68) | 239.84(149.84, 383.9) | 4145.99 | 7.51(6.86) | 182.01(122.88) |
| spermatocele | 22 | 660.92(372.64, 1172.21) | 660.7(374.24, 1166.43) | 7708.26 | 8.46(7.74) | 351.91(217.87) |
| testicular cyst | 21 | 133.66(84.02, 212.63) | 133.62(83.48, 213.88) | 2346.52 | 6.83(6.18) | 113.58(77.02) |
| penile vascular disorder | 17 | 170.22(100.54, 288.2) | 170.18(100.25, 288.89) | 2330.86 | 7.12(6.39) | 138.92(89.42) |
| breast disorder male | 17 | 138.77(82.71, 232.83) | 138.73(83.34, 230.93) | 1962.06 | 6.87(6.15) | 117.25(76.05) |
| hypospermia | 16 | 750.98(375.54, 1501.75) | 750.8(378.09, 1490.91) | 5990.42 | 8.55(7.71) | 375.9(210.49) |
| feminisation acquired | 16 | 387.6(212, 708.66) | 387.51(211.06, 711.48) | 4068.39 | 8(7.2) | 255.93(154.48) |
| prostatic calcification | 16 | 130.6(76.8, 222.1) | 130.57(76.92, 221.65) | 1752.52 | 6.8(6.06) | 111.38(71.43) |
| genital disorder | 13 | 157.46(86.59, 286.31) | 157.43(85.75, 289.04) | 1670.38 | 7.03(6.2) | 130.31(79.01) |
| testis discomfort | 13 | 131.92(73.17, 237.86) | 131.9(73.26, 237.47) | 1436.42 | 6.81(6) | 112.34(68.6) |
| penile discomfort | 12 | 121.77(66.17, 224.12) | 121.75(66.31, 223.54) | 1236.59 | 6.71(5.87) | 104.9(62.97) |
| teratospermia | 10 | 208.59(103.51, 420.32) | 208.56(102.99, 422.35) | 1616.56 | 7.35(6.41) | 163.43(90.94) |
| prostate tenderness | 9 | 168.95(81.98, 348.2) | 168.93(81.8, 348.86) | 1226.47 | 7.11(6.13) | 138.09(75.4) |
| testicular microlithiasis | 6 | 250.29(99.35, 630.55) | 250.27(99.62, 628.75) | 1117.22 | 7.55(6.34) | 187.95(86.75) |
| prostatic atrophy | 5 | 250.29(90.96, 688.68) | 250.27(90.32, 693.49) | 931.02 | 7.55(6.25) | 187.95(80.58) |
| epididymal disorder | 4 | 176.67(59.44, 525.06) | 176.66(58.95, 529.44) | 565.58 | 7.16(5.76) | 143.2(57.56) |
| epididymal tenderness | 3 | 450.5(107.66, 1885.15) | 450.48(107.72, 1883.93) | 840.9 | 8.14(6.44) | 281.93(85.11) |
| oligoasthenozoospermia | 3 | 160.89(46.24, 559.89) | 160.89(45.89, 564.04) | 392.56 | 7.05(5.49) | 132.67(46.73) |
| **renal and urinary disorders** |  |  |  |  |  |  |
| terminal dribbling | 13 | 278.92(147.57, 527.2) | 278.87(148.94, 522.15) | 2624.52 | 7.67(6.81) | 203.61(119.52) |
| post micturition dribble | 6 | 136.52(57.2, 325.83) | 136.51(57.63, 323.37) | 682.93 | 6.85(5.7) | 115.66(55.86) |
| **endocrine disorders** |  |  |  |  |  |  |
| hypogonadism | 302 | 365.59(318.58, 419.54) | 363.95(317.29, 417.47) | 73623.19 | 7.94(7.75) | 245.45(218.75) |
| post 5-alpha-reductase inhibitor syndrome | 166 | 10411.85(5795.02, 18706.87) | 10386.07(5768.81, 18698.91) | 116208.64 | 9.45(9.15) | 701.12(429.4) |
| hypogonadism male | 79 | 418.19(317.58, 550.67) | 417.7(317.46, 549.59) | 21101.15 | 8.07(7.7) | 268.74(213.47) |
| androgen deficiency | 77 | 238.18(184.31, 307.8) | 237.91(184.4, 306.95) | 13794.31 | 7.5(7.14) | 180.9(145.97) |
| testicular failure | 72 | 1061.09(741.19, 1519.07) | 1059.95(744.85, 1508.36) | 31583.88 | 8.78(8.36) | 440.08(325.95) |
| secondary hypogonadism | 42 | 137.19(98.73, 190.63) | 137.1(98.25, 191.31) | 4798.42 | 6.86(6.39) | 116.09(88.15) |
| oestrogenic effect | 4 | 333.71(102.76, 1083.67) | 333.69(102.95, 1081.62) | 918.53 | 7.85(6.38) | 231.32(86.34) |
| **psychiatric disorders** |  |  |  |  |  |  |
| libido decreased | 1476 | 119.46(113.02, 126.26) | 116.85(110.18, 123.93) | 146728.83 | 6.66(6.58) | 101.25(96.66) |
| loss of libido | 1301 | 160.75(151.35, 170.73) | 157.65(148.65, 167.2) | 167389.74 | 7.03(6.94) | 130.46(124.05) |
| orgasm abnormal | 109 | 135.26(110.3, 165.88) | 135.04(111, 164.28) | 12291.97 | 6.84(6.55) | 114.61(96.62) |
| male orgasmic disorder | 108 | 240.29(193.46, 298.45) | 239.9(193.37, 297.62) | 19472.09 | 7.51(7.21) | 182.05(151.85) |
| premature ejaculation | 83 | 124.29(98.52, 156.8) | 124.14(98.12, 157.06) | 8699.7 | 6.74(6.41) | 106.67(87.82) |
| orgasmic sensation decreased | 74 | 228.89(176.43, 296.96) | 228.64(177.21, 294.99) | 12856.54 | 7.46(7.09) | 175.5(141.15) |
| psychogenic erectile dysfunction | 8 | 600.71(237.07, 1522.11) | 600.64(239.08, 1508.99) | 2660.63 | 8.38(7.24) | 334.13(153.48) |
| catastrophic reaction | 8 | 150.18(70.29, 320.85) | 150.16(69.92, 322.5) | 987.78 | 6.97(5.94) | 125.3(66.39) |
| psychophysiologic insomnia | 5 | 288.79(102.95, 810.09) | 288.77(102.19, 816.01) | 1035.57 | 7.71(6.38) | 208.83(88.1) |
| thermophobia | 3 | 150.17(43.47, 518.73) | 150.16(43.68, 516.2) | 370.42 | 6.97(5.41) | 125.3(44.41) |
| **congenital, familial and genetic disorders** |  |  |  |  |  |  |
| reproductive tract hypoplasia, male | 5 | 234.64(85.96, 640.53) | 234.63(86.35, 637.54) | 886.21 | 7.48(6.18) | 179(77.26) |
| androgen insensitivity syndrome | 4 | 375.42(113.04, 1246.79) | 375.4(113.57, 1240.9) | 995.74 | 7.97(6.48) | 250.6(91.79) |
| pseudohermaphroditism male | 3 | 2252.5(234.29, 21655.66) | 2252.4(236.45, 21455.83) | 1687.8 | 9.14(7.24) | 563.85(84.87) |
| **surgical and medical procedures** |  |  |  |  |  |  |
| hair transplant | 49 | 2832.01(1536.43, 5220.08) | 2829.94(1541.35, 5195.83) | 29054.83 | 9.21(8.68) | 594.16(356.19) |
| vasectomy | 31 | 173.77(117.57, 256.84) | 173.69(117.36, 257.05) | 4322.65 | 7.14(6.6) | 141.25(101.86) |
| radical prostatectomy | 15 | 154.31(88.52, 268.98) | 154.27(89.11, 267.07) | 1894.86 | 7(6.23) | 128.15(80.5) |
| varicocele repair | 8 | 2002.37(531.2, 7547.99) | 2002.13(528.04, 7591.37) | 4363.93 | 9.09(7.85) | 546.76(180.14) |
| radiotherapy to prostate | 8 | 333.73(145.1, 767.55) | 333.69(146.5, 760.07) | 1837.06 | 7.85(6.77) | 231.32(115.23) |
| scrotal operation | 6 | 409.56(151.46, 1107.5) | 409.53(150.72, 1112.77) | 1582.17 | 8.05(6.79) | 265.34(115.43) |
| penile prosthesis insertion | 6 | 128.72(54.14, 306.04) | 128.71(54.33, 304.89) | 649.04 | 6.78(5.63) | 110.02(53.3) |
| rectal fistula repair | 3 | 173.27(49.37, 608.06) | 173.26(49.42, 607.4) | 417.46 | 7.14(5.57) | 140.96(49.31) |
| uvulopalatopharyngoplasty | 3 | 125.14(36.86, 424.84) | 125.13(37.12, 421.81) | 316.65 | 6.75(5.21) | 107.4(38.62) |
| **nervous system disorders** |  |  |  |  |  |  |
| pudendal canal syndrome | 12 | 214.55(112.95, 407.55) | 214.51(112.34, 409.59) | 1983.51 | 7.38(6.51) | 167.07(97.66) |

ROR, reporting odds ratio; CI, confidence interval; PRR, proportional reporting ratio; chisq, chi-squared; IC, information component; EBGM, empirical

Table2
| PTs and categories | Case Reports | ROR(95% CI) | PRR(95% CI) | chisq | IC(IC025) | EBGM(EBGM05) |
| --- | --- | --- | --- | --- | --- | --- |
| reproductive system and breast disorders |  |  |  |  |  |  |
| erectile dysfunction | 3377 | 144.15(138.85, 149.66) | 136.94(131.68, 142.41) | 385588.1 | 6.86(6.8) | 115.97(112.39) |
| sexual dysfunction | 2160 | 220.84(210.4, 231.79) | 213.75(205.53, 222.3) | 356099.9 | 7.38(7.31) | 166.6(159.99) |
| ejaculation disorder | 652 | 243.38(222.76, 265.92) | 241.02(222.85, 260.68) | 117980.6 | 7.51(7.39) | 182.7(169.65) |
| testicular pain | 618 | 223(203.8, 244) | 220.95(200.32, 243.7) | 104552.7 | 7.42(7.29) | 170.94(158.54) |
| ejaculation failure | 487 | 279.44(251.79, 310.14) | 277.42(251.52, 305.98) | 97945.39 | 7.66(7.52) | 202.84(185.9) |
| penile size reduced | 398 | 662.12(578.62, 757.67) | 658.19(573.81, 754.98) | 139167.4 | 8.46(8.28) | 351.19(313.73) |
| genital hypoaesthesia | 235 | 337.89(289.65, 394.18) | 336.71(287.84, 393.87) | 54305.2 | 7.86(7.65) | 232.77(204.62) |
| organic erectile dysfunction | 218 | 1094.72(889.08, 1347.92) | 1091.16(879.54, 1353.7) | 96782.38 | 8.8(8.56) | 445.36(374.2) |
| peyronie's disease | 215 | 286.11(244.49, 334.82) | 285.2(243.81, 333.62) | 44127.17 | 7.69(7.48) | 206.96(181.45) |
| testicular atrophy | 213 | 301.56(257.2, 353.56) | 300.6(256.98, 351.63) | 45419.19 | 7.75(7.53) | 214.94(188.15) |
| testicular disorder | 158 | 157.28(132.48, 186.73) | 156.91(131.53, 187.18) | 20246.36 | 7.02(6.78) | 129.96(112.58) |
| painful ejaculation | 96 | 707.65(535.46, 935.2) | 706.64(537.07, 929.76) | 34847.63 | 8.51(8.16) | 364.51(288.66) |
| varicocele | 89 | 461.45(354.37, 600.89) | 460.84(357.18, 594.58) | 25304.84 | 8.16(7.81) | 285.94(229.26) |
| male sexual dysfunction | 74 | 136.32(106.41, 174.65) | 136.17(105.54, 175.69) | 8405.1 | 6.85(6.5) | 115.42(93.81) |
| prostatic pain | 68 | 173.83(133.52, 226.32) | 173.65(134.59, 224.04) | 9480.26 | 7.14(6.77) | 141.22(113.25) |
| infertility male | 64 | 136.25(104.39, 177.84) | 136.12(103.46, 179.1) | 7266.87 | 6.85(6.47) | 115.38(92.33) |
| penile curvature | 63 | 328.78(244.5, 442.11) | 328.48(244.81, 440.75) | 14308.32 | 7.84(7.43) | 228.81(178.59) |
| male genital atrophy | 52 | 1116.34(727.21, 1713.69) | 1115.47(724.75, 1716.82) | 23293.43 | 8.81(8.32) | 449.35(313.93) |
| genital atrophy | 49 | 944(619.84, 1437.69) | 943.31(625.03, 1423.67) | 20441.53 | 8.71(8.21) | 418.62(294.41) |
| epididymal cyst | 49 | 800.35(535.17, 1196.92) | 799.77(529.92, 1207.04) | 18928.1 | 8.6(8.11) | 387.77(276.9) |
| genital paraesthesia | 49 | 230.1(167.07, 316.91) | 229.93(168.04, 314.62) | 8550.4 | 7.46(7.02) | 176.26(134.84) |
| scrotal disorder | 38 | 115.57(82.13, 162.64) | 115.51(82.78, 161.19) | 3738.49 | 6.65(6.16) | 100.24(75.32) |
| male reproductive tract disorder | 26 | 330.99(208.65, 525.06) | 330.86(206.71, 529.59) | 5935.04 | 7.85(7.22) | 229.96(156.31) |
| oligospermia | 23 | 239.92(150.02, 383.68) | 239.84(149.84, 383.9) | 4145.99 | 7.51(6.86) | 182.01(122.88) |
| spermatocele | 22 | 660.92(372.64, 1172.21) | 660.7(374.24, 1166.43) | 7708.26 | 8.46(7.74) | 351.91(217.87) |
| testicular cyst | 21 | 133.66(84.02, 212.63) | 133.62(83.48, 213.88) | 2346.52 | 6.83(6.18) | 113.58(77.02) |
| penile vascular disorder | 17 | 170.22(100.54, 288.2) | 170.18(100.25, 288.89) | 2330.86 | 7.12(6.39) | 138.92(89.42) |
| breast disorder male | 17 | 138.77(82.71, 232.83) | 138.73(83.34, 230.93) | 1962.06 | 6.87(6.15) | 117.25(76.05) |
| hypospermia | 16 | 750.98(375.54, 1501.75) | 750.8(378.09, 1490.91) | 5990.42 | 8.55(7.71) | 375.9(210.49) |
| feminisation acquired | 16 | 387.6(212, 708.66) | 387.51(211.06, 711.48) | 4068.39 | 8(7.2) | 255.93(154.48) |
| prostatic calcification | 16 | 130.6(76.8, 222.1) | 130.57(76.92, 221.65) | 1752.52 | 6.8(6.06) | 111.38(71.43) |
| genital disorder | 13 | 157.46(86.59, 286.31) | 157.43(85.75, 289.04) | 1670.38 | 7.03(6.2) | 130.31(79.01) |
| testis discomfort | 13 | 131.92(73.17, 237.86) | 131.9(73.26, 237.47) | 1436.42 | 6.81(6) | 112.34(68.6) |
| penile discomfort | 12 | 121.77(66.17, 224.12) | 121.75(66.31, 223.54) | 1236.59 | 6.71(5.87) | 104.9(62.97) |
| teratospermia | 10 | 208.59(103.51, 420.32) | 208.56(102.99, 422.35) | 1616.56 | 7.35(6.41) | 163.43(90.94) |
| prostate tenderness | 9 | 168.95(81.98, 348.2) | 168.93(81.8, 348.86) | 1226.47 | 7.11(6.13) | 138.09(75.4) |
| testicular microlithiasis | 6 | 250.29(99.35, 630.55) | 250.27(99.62, 628.75) | 1117.22 | 7.55(6.34) | 187.95(86.75) |
| prostatic atrophy | 5 | 250.29(90.96, 688.68) | 250.27(90.32, 693.49) | 931.02 | 7.55(6.25) | 187.95(80.58) |
| epididymal disorder | 4 | 176.67(59.44, 525.06) | 176.66(58.95, 529.44) | 565.58 | 7.16(5.76) | 143.2(57.56) |
| epididymal tenderness | 3 | 450.5(107.66, 1885.15) | 450.48(107.72, 1883.93) | 840.9 | 8.14(6.44) | 281.93(85.11) |
| oligoasthenozoospermia | 3 | 160.89(46.24, 559.89) | 160.89(45.89, 564.04) | 392.56 | 7.05(5.49) | 132.67(46.73) |
| renal and urinary disorders |  |  |  |  |  |  |
| terminal dribbling | 13 | 278.92(147.57, 527.2) | 278.87(148.94, 522.15) | 2624.52 | 7.67(6.81) | 203.61(119.52) |
| post micturition dribble | 6 | 136.52(57.2, 325.83) | 136.51(57.63, 323.37) | 682.93 | 6.85(5.7) | 115.66(55.86) |
| endocrine disorders |  |  |  |  |  |  |
| hypogonadism | 302 | 365.59(318.58, 419.54) | 363.95(317.29, 417.47) | 73623.19 | 7.94(7.75) | 245.45(218.75) |
| post 5-alpha-reductase inhibitor syndrome | 166 | 10411.85(5795.02, 18706.87) | 10386.07(5768.81, 18698.91) | 116208.6 | 9.45(9.15) | 701.12(429.4) |
| hypogonadism male | 79 | 418.19(317.58, 550.67) | 417.7(317.46, 549.59) | 21101.15 | 8.07(7.7) | 268.74(213.47) |
| androgen deficiency | 77 | 238.18(184.31, 307.8) | 237.91(184.4, 306.95) | 13794.31 | 7.5(7.14) | 180.9(145.97) |
| testicular failure | 72 | 1061.09(741.19, 1519.07) | 1059.95(744.85, 1508.36) | 31583.88 | 8.78(8.36) | 440.08(325.95) |
| secondary hypogonadism | 42 | 137.19(98.73, 190.63) | 137.1(98.25, 191.31) | 4798.42 | 6.86(6.39) | 116.09(88.15) |
| oestrogenic effect | 4 | 333.71(102.76, 1083.67) | 333.69(102.95, 1081.62) | 918.53 | 7.85(6.38) | 231.32(86.34) |
| psychiatric disorders |  |  |  |  |  |  |
| libido decreased | 1476 | 119.46(113.02, 126.26) | 116.85(110.18, 123.93) | 146728.8 | 6.66(6.58) | 101.25(96.66) |
| loss of libido | 1301 | 160.75(151.35, 170.73) | 157.65(148.65, 167.2) | 167389.7 | 7.03(6.94) | 130.46(124.05) |
| orgasm abnormal | 109 | 135.26(110.3, 165.88) | 135.04(111, 164.28) | 12291.97 | 6.84(6.55) | 114.61(96.62) |
| male orgasmic disorder | 108 | 240.29(193.46, 298.45) | 239.9(193.37, 297.62) | 19472.09 | 7.51(7.21) | 182.05(151.85) |
| premature ejaculation | 83 | 124.29(98.52, 156.8) | 124.14(98.12, 157.06) | 8699.7 | 6.74(6.41) | 106.67(87.82) |
| orgasmic sensation decreased | 74 | 228.89(176.43, 296.96) | 228.64(177.21, 294.99) | 12856.54 | 7.46(7.09) | 175.5(141.15) |
| psychogenic erectile dysfunction | 8 | 600.71(237.07, 1522.11) | 600.64(239.08, 1508.99) | 2660.63 | 8.38(7.24) | 334.13(153.48) |
| catastrophic reaction | 8 | 150.18(70.29, 320.85) | 150.16(69.92, 322.5) | 987.78 | 6.97(5.94) | 125.3(66.39) |
| psychophysiologic insomnia | 5 | 288.79(102.95, 810.09) | 288.77(102.19, 816.01) | 1035.57 | 7.71(6.38) | 208.83(88.1) |
| thermophobia | 3 | 150.17(43.47, 518.73) | 150.16(43.68, 516.2) | 370.42 | 6.97(5.41) | 125.3(44.41) |
| congenital, familial and genetic disorders |  |  |  |  |  |  |
| reproductive tract hypoplasia, male | 5 | 234.64(85.96, 640.53) | 234.63(86.35, 637.54) | 886.21 | 7.48(6.18) | 179(77.26) |
| androgen insensitivity syndrome | 4 | 375.42(113.04, 1246.79) | 375.4(113.57, 1240.9) | 995.74 | 7.97(6.48) | 250.6(91.79) |
| pseudohermaphroditism male | 3 | 2252.5(234.29, 21655.66) | 2252.4(236.45, 21455.83) | 1687.8 | 9.14(7.24) | 563.85(84.87) |
| surgical and medical procedures |  |  |  |  |  |  |
| hair transplant | 49 | 2832.01(1536.43, 5220.08) | 2829.94(1541.35, 5195.83) | 29054.83 | 9.21(8.68) | 594.16(356.19) |
| vasectomy | 31 | 173.77(117.57, 256.84) | 173.69(117.36, 257.05) | 4322.65 | 7.14(6.6) | 141.25(101.86) |
| radical prostatectomy | 15 | 154.31(88.52, 268.98) | 154.27(89.11, 267.07) | 1894.86 | 7(6.23) | 128.15(80.5) |
| varicocele repair | 8 | 2002.37(531.2, 7547.99) | 2002.13(528.04, 7591.37) | 4363.93 | 9.09(7.85) | 546.76(180.14) |
| radiotherapy to prostate | 8 | 333.73(145.1, 767.55) | 333.69(146.5, 760.07) | 1837.06 | 7.85(6.77) | 231.32(115.23) |
| scrotal operation | 6 | 409.56(151.46, 1107.5) | 409.53(150.72, 1112.77) | 1582.17 | 8.05(6.79) | 265.34(115.43) |
| penile prosthesis insertion | 6 | 128.72(54.14, 306.04) | 128.71(54.33, 304.89) | 649.04 | 6.78(5.63) | 110.02(53.3) |
| rectal fistula repair | 3 | 173.27(49.37, 608.06) | 173.26(49.42, 607.4) | 417.46 | 7.14(5.57) | 140.96(49.31) |
| uvulopalatopharyngoplasty | 3 | 125.14(36.86, 424.84) | 125.13(37.12, 421.81) | 316.65 | 6.75(5.21) | 107.4(38.62) |
| nervous system disorders |  |  |  |  |  |  |
| pudendal canal syndrome | 12 | 214.55(112.95, 407.55) | 214.51(112.34, 409.59) | 1983.51 | 7.38(6.51) | 167.07(97.66) |

When the number of cases surpassed 100, this signaled a strong indication of an adverse event[29]. After excluding PT as a signal of drug-independence of finasteride, we screened PT entries with more than 100 cases sorting by the number of cases in a descending fashion, reproductive system and breast disorder-related PT: erectile dysfunction (n= 3377), sexual dysfunction (n= 2160), ejaculatory dysfunction (n= 652), testicular pain (n= 618), ejaculatory failure (n= 487), penile size reduction (n= 398), hypesthesia of the genitals (n= 235), organic erectile dysfunction (n= 218), Peyronie’s disease (n= 215), testicular atrophy (n= 213), and testicular disorder (n= 158); and psychiatric disorders-related PTs: decreased sexual desire (n= 1476), loss of libido (n= 1301), orgasm abnormal (n= 109), male orgasmic disorder (n= 108); and endocrine disorders-related PT: hypogonadism (n= 302), post-5α reductase inhibitor syndrome (n= 166). It is worth noting that endocrine system disorders are an adverse reaction disease system not mentioned in the specification, suggesting that we should not ignore the occurrence of adverse reactions in such systems.

In order to improve the stability of the calculations with fewer cases[30], we also a1nalyzed the IC values and found that male pseudohermaphroditism male (n= 3, IC= 9.14), varicocele repair (n= 8, IC= 9.09), psychogenic erectile dysfunction (n= 8, IC= 8.38), epididymal tenderness (n= 3, IC= 8.14), scrotal operation (n= 6, IC= 8.05), and androgen insensitivity syndrome (n=4, IC=7.97) were among the unexpected signals with higher IC values. This implies a robust correlation with finasteride dosage.

To classify the early warning signals more carefully[31], we performed a descending order ranking of the BCPNN algorithm, and after excluding drug-unrelated signals, the top 10 finasteride PTs were post-5α reductase inhibitor syndrome (EBGM=701.12), hair transplant (EBGM=594.16), pseudohermaphroditism male (EBGM=563.85), varicocele repair (EBGM=546.76), male genital atrophy (EBGM=449.35), organic erectile dysfunction (EBGM=445.36), testicular failure (EBGM=440.08), genital atrophy (EBGM=418.62), epididymal cysts (EBGM=387.77), and hypospermia (EBGM= 375.9).

In summary, we found that ejaculation disorder, sexual dysfunction, ejaculation disorder, and epididymal tenderness were consistent with the drug insert and dosing warnings. Unexpectedly post-5α reductase inhibitor syndrome, hair transplant, male pseudohermaphroditism male, Peyronie’s disease, male genital atrophy, epididymal cysts, testicular failure, varicocele repair, and androgen insensitivity syndrome were absent from the drug insert and require additional investigation.

### Time to Onset Analysis

Out of all adverse events reported, 7019 included the onset time, with a median onset time of 61 days (interquartile range 6.00-463.75). After excluding inaccurate, missing, or unknown gender time of onset reports, a total of 7019 finasteride-administered AEs reported time of onset. In Fig 3, it was shown that the time of onset in males (n=1208) versus females (n=47) was predominantly within one month after initiation of finasteride medication. Interestingly, after one year of finasteride treatment, adverse events could still occur in males (n=879) and females (n=11). This indicates the importance of ongoing patient monitoring for potential adverse events even beyond one year of finasteride treatment.

**Fig 3.**
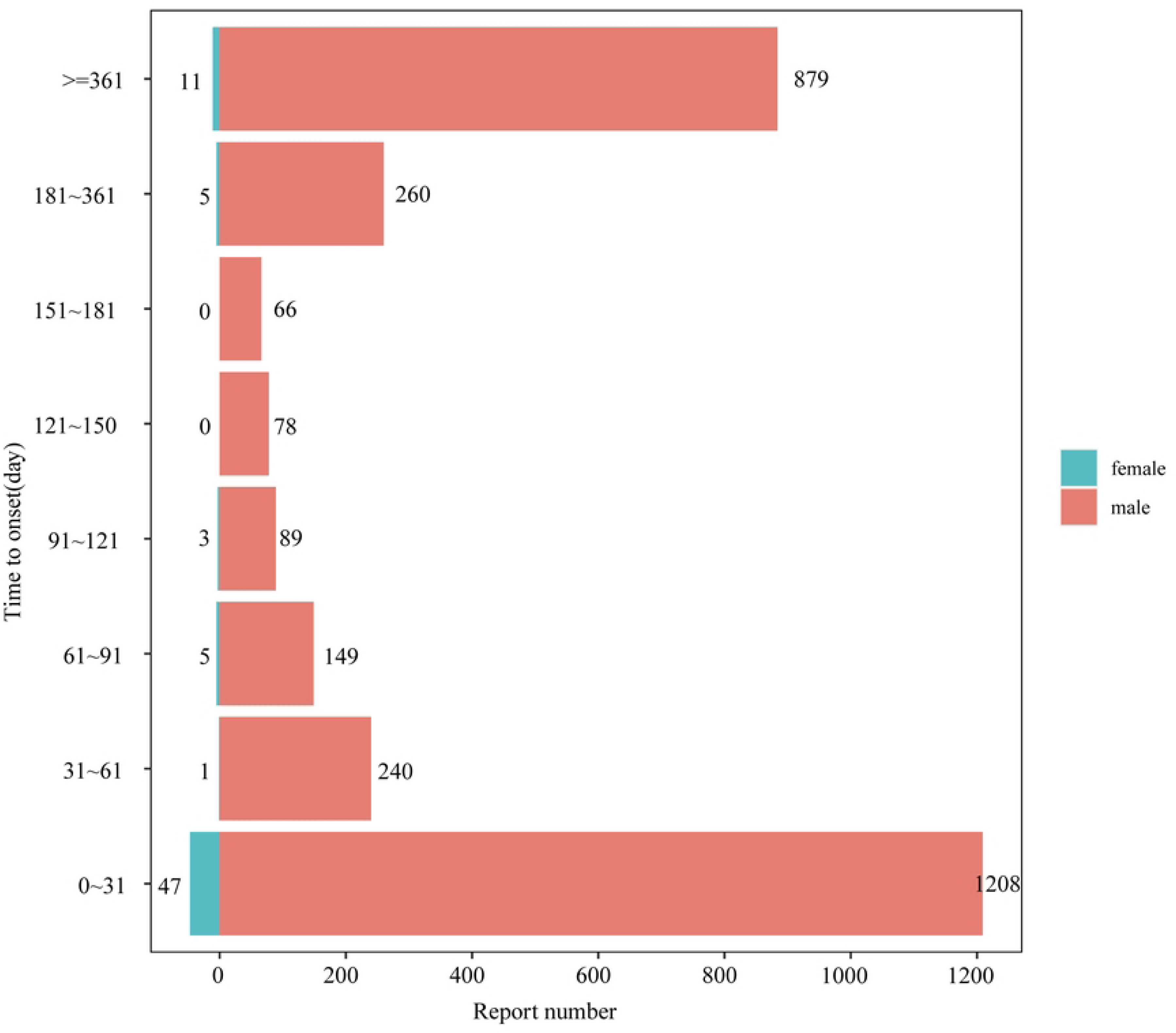
Time to onset of AEs in male and female patients receiving finasteride therapy. AEs: adverse events.

### Subgroup analysis

#### Gender in different PT groups

To investigate the impact of gender on adverse reactions to finasteride treatment, we identified PTs for 102 male and 7 female adverse reactions using four statistical methods, with the results detailed in S4-S5 Tables. Following subgroup analyses where we excluded indications mentioned in adverse reaction reports, depression and anxiety emerged as frequent PTs for both men and women.

The ‘volcano plot’ in Fig 4 visualizes gender-specific AE signals following finasteride treatment. Every data point in the figure represents an adverse event associated with finasteride treatment, and we have labelled statistically significant AEs. Blue points denote potential adverse event signals in female patients, whereas red points represent potential adverse event signals in male patients. The uterine cervix stenosis was a more frequent AE in females compared to males, whereas post micturition dribble was a frequent AE in males. The results above provide gender-specific insights into potential adverse event signals related to finasteride therapy, emphasizing that reported adverse events vary between men and women and necessitate distinct considerations in clinical management.

**Fig 4.**
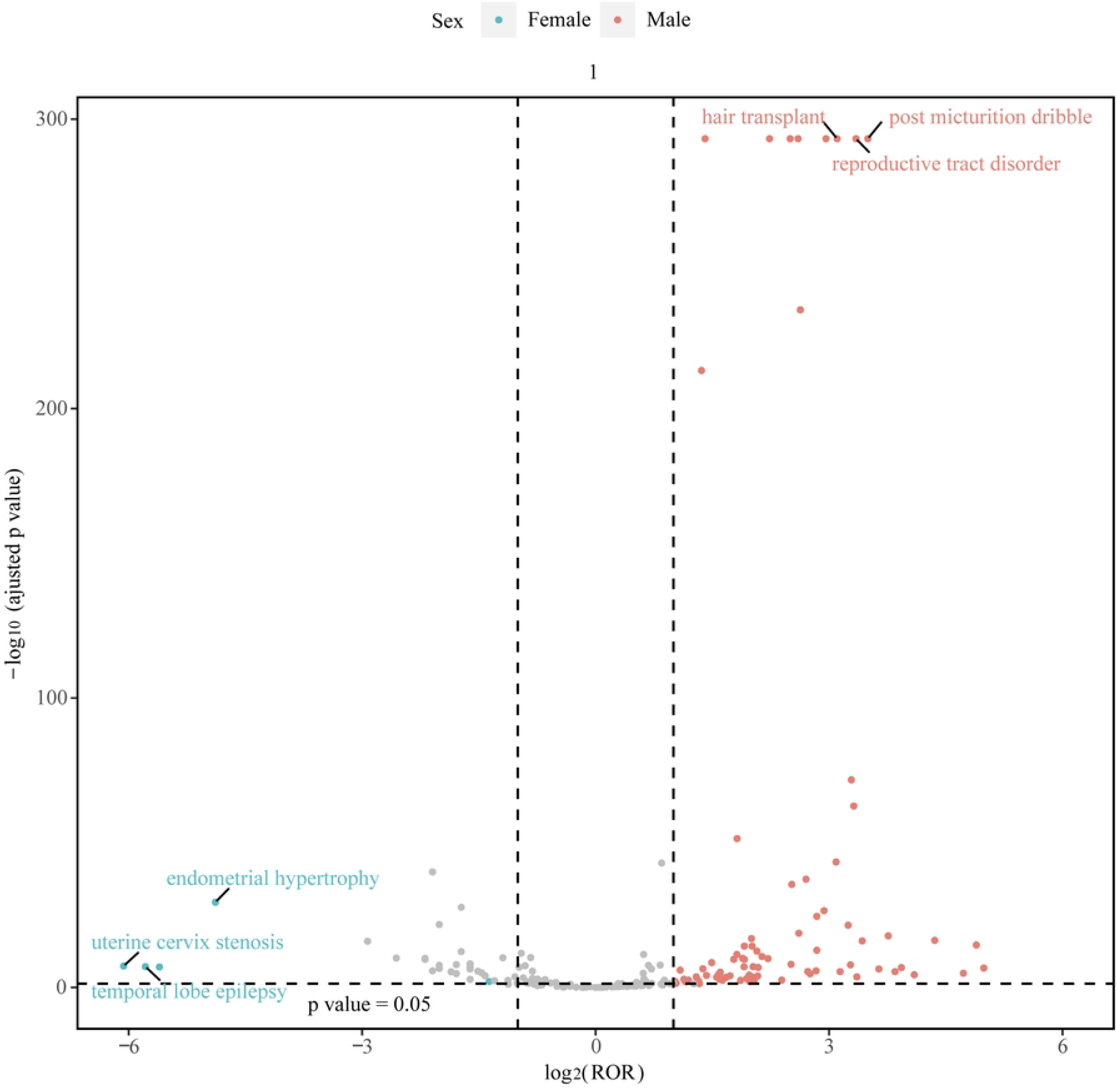
Gender-differentiated risk signal volcano plot for finasteride.

The horizontal coordinate shows the log2 PRR value and the vertical coordinate indicates the adjusted p-value after −log10 conversion PRR, proportional reporting ratio.

#### Age in different PT groups

Age is a risk factor for androgenetic alopecia[32]. The incidence of AGA increases with age[33]. In order to minimize the confounding effect of age on the study of adverse effects, we performed an age-stratified analysis.

To analyze whether age affects adverse reactions to finasteride medication, we used four statistical methods to determine the PT of 17 adverse reactions in patients aged less than 18 years, 157 in patients aged between 18 and 65 years, and 133 in patients aged greater than or equal to 65 years, with the results presented in S6-S8 Tables. We arranged the number of cases in descending order and excluded the indications specified in the adverse reaction reports. The top three PTs in patients younger than 18 years were depression (n=11), erectile dysfunction (n=10), and cryptorchism (n=8). The top three PTs in patients between the ages of 18 and 65 were erectile dysfunction (n=1435), depression (n=979), and anxiety (n=704). The top three PTs in patients older than 65 years were depression (n=102), gynaecomastia (n=73), and erectile dysfunctio (n=70). Erectile dysfunction, libido decreased, depression, and suicidal ideation, etc. were common AEs in patients of different ages.

As most of the cases were less than 50 after subgroup analysis, we used IC values for descending order. The top three PTs in patients younger than 18 years were scrotal disorder (IC= 10.92), libido decreased (IC= 9.38), and loss of libido (IC= 9.01). The top three PTs in patients aged 18 to 65 years were post 5-alpha-reductase inhibitor syndrome (IC= 9.55), male genital atrophy (IC= 9.32) and genital atrophy (IC= 8.89). The top three PTs among patients aged greater than or equal to 65 years were prostate tenderness (IC= 9.98), penile size reduction (IC= 9.57) and penile exfoliation (IC= 9.33). Finally, of all the PTs, the highest PTs with IC values for patients younger than 18 years of age were found in reproductive system and breast disorders, the highest PTs with IC values for patients between 18 and 65 years of age were found in endocrine disorders, and the highest PTs with IC values for patients older than or equal to 65 years of age were found in reproductive system and breast disorders. Erectile dysfunction, libido decreased, depression, suicidal ideation, psychotic disorder and disturbance in attention were common PTs in patients of different ages. The above suggests that different age groups have different adverse events and focus on different SOCs.

## Discussion

To our knowledge, earlier studies on finasteride safety have primarily been confined to clinical trials or concentrated on particular adverse events, such as sexual dysfunction, and have mainly investigated potential safety concerns[34–36], whereas our study discusses this more fully and extensively. Although in a large real-world drug study in 2017 it comprehensively and systematically summarized global reports of finasteride-associated sexual dysfunction in FAERS, we offered a more precise and comprehensive description and identification of adverse events associated with finasteride. Although the incidence of AEs reported in clinical trials was low, the risk of occurrence of finasteride-associated AEs still exists, necessitating further studies and additions to the literature.

With regard to ‘reproductive system and breast disorders’, sexual dysfunction is the most significant adverse effect of finasteride treatment in young men with AGA, manifested by loss of libido and erectile dysfunction[37]. Moreover, the distribution of medication-related AEs indicated that the highest number of cases (3146 27.22%) were in the age group of 18-45 years, and in the younger age group even a low dose of finasteride (1mg) may lead to persistent sexual dysfunction in young males, which may increase the risk of suicide [38]and persist after discontinuation of the medication[8]. This is probably due to the fact that the younger patients have a more stressful life and are more sensitive to hormonal changes than the older ones[39]. In addition, in the PT analysis we observed ‘sexual dysfunction’ and ‘catastrophic reaction’. This may be due to the fact that younger patients have more stressful lives, are more sensitive to hormonal changes than older adults, are more prone to symptoms, and are more susceptible to the adverse effects of finasteride. These two adverse effects further confirm the existing knowledge about the potential adverse consequences of finasteride on sexual functioning and the increased risk of suicide[40]. It was found that an increased risk of suicide may exist with finasteride in the treatment of men with a history of psychological disorders[41]. Surprisingly, suicidality and depression in patients with androgenetic alopecia may be the side effects of finasteride --sexual dysfunction (erectile dysfunction, hyperectile dysfunction, neurological deficits) caused by finasteride[42]. Therefore, comprehensive and precise clinical screening for the drug and close monitoring of patients for post-drug adverse effects are warranted.

With regard to the ‘psychiatric disorders’, psychological distress in patients with androgenetic alopecia has become a significant motivator in the development of effective treatments and the reduction of adverse effects of medication[43]. There is a risk of suicide, depression, and anxiety in patients younger than 40 years of age treated with finasteride for alopecia areata[44], which may be related to the involvement of 5 alpha-reductase inhibitors in the central nervous system’s synthesis of neuroactive steroidal organisms[45]. Neurosteroids are important physiological regulators of neural function in the adult brain and are involved in the control of the neuroendocrine system of reproduction[46]. A small human study observed depressive symptoms associated with reduced levels of neurosteroids in the cerebrospinal fluid of men who were taking finasteride[47]. This observation was supported by animal studies[48,49]. A study has been conducted by evaluating FMRI and HPLC-mass spectrometry analyses of neurosteroid levels in cerebrospinal fluid, pointing to the fact that finasteride disrupts neurotransmitters and chemical messengers[50]. These findings reinforce that anxiety and depression induced by finasteride use cannot be ignored. Lastly, increased levels of corticosterone are one of the potential pathways through which finasteride may induce anxiety and depression, with plasma cortisol elevated in rats following short-term finasteride administration[51]. Emergingevidence also suggests that finasteride inhibits glioblastoma proliferation[52]. These findings highlight the potential relevance of finasteride to neuroscience and the need to closely monitor neurological psychiatric problems in patients receiving finasteride.

With regard to ‘renal and urinary disorders’, the only pt’s with positive signals were post micturition dribble and terminal dribbling, the former of which is the most frequent adverse reaction in men. Traish suggests that the symptoms are due to a novel tissue-specific androgen deficiency that develops when finasteride inhibits the enzyme 5-alpha reductase, which reduces the biosynthesis of 5alpha-dihydrotestosterone[53]. The symptoms of post micturition dribble have been shown to be associated with the development of a new form of androgen deficiency in men. In addition, finasteride has been widely used in the treatment of a variety of testosterone-related disorders, and new evidence suggests that finasteride has a protective effect on testosterone-induced calcium oxalate crystals, which is effective in preventing the formation of calcium oxalate-type renal stones and neutralising the calcium oxalate-promoting effects of testosterone[54]. Future studies could explore the relationship between finasteride and testosterone levels, and reduce the prevalence of renal stones in patients with high levels of the testosterone.

At the PT level, we discovered that while prostate pain, prostate calcification, and prostate atrophy were notable in disproportionate assessments and identified as adverse reactions by certain studies, they were regarded as treatment indications in Table 1, consistent with findings from another research. Although finasteride reduces the risk of prostate cancer, it increases the malignancy of high-grade prostate cancer[55]. To ensure the reliability of our results, we omitted adverse reactions listed as indications in Table 2.

In addition to the expected AEs associated with finasteride use, our study identified a number of unintended AEs not mentioned in the specification that need to be further analyzed and evaluated, including ‘Peyronie’s disease’, ‘post-5α reductase inhibitor syndrome’, ‘catastrophic reaction’ and “thermophobia”. Peyronie’s disease is a male condition characterized by penile curvature, pain, penile shortening and erectile dysfunction[56]. This is an unintended AE associated with androgenetic alopecia treated with finasteride (176/210 83.8%) [57]and the results of our study are consistent with the above studies. In addition, a case report found that the pathogenesis of finasteride-treated androgenetic alopecia is the same as that of Peyronie’s disease, in that inflammation affects the penile leukomalacia, leading to the formation of inelastic microtissue that deforms the penis[58,59]. However, the relationship needs to be further determined[57]. Another specification unmentionable AE that appeared in our analysis was the post 5-alpha-reductase inhibitor syndrome, 5alpha-reductase inhibitors are drugs used to treat androgen-dependent diseases. Long-term use of 5-alpha-reductase inhibitors in patients with androgenetic alopecia leads to persistent side effects known as post-finasteride syndrome (PFS) [60,61]. PFS is also characterized by symptoms of penile shortening and penile curvature[62]. Not only this, but also persistent or irreversible sexual, neurological, physical and psychiatric side effects[63]. Leliefeld et al. have demonstrated that finasteride, a 5α-reductase inhibitor, inhibits the conversion of testosterone to 5α dihydrotestosterone (DHT) in the prostate and scalp and impairs the levels of other neurosteroids in the brain, demonstrating that sustained changes in androgen receptor and neurosteroid levels are associated with PFS[64]. However, previous studies have been inconclusive as to whether or not PFS exists[65]. Finally, our study reinforces the need for additional clinical research to assess the balance between the health risks and benefits of the medication. Careful consideration is required when using finasteride and developing potential new therapeutic options for treating these two conditions.

There is no mention of the adverse reaction in the insert: male breast cancer in the FAERS database or in the previous literature. As for the rare adverse reaction in the specification: hypersensitivity reaction, there was one case report of a patient with prostatic hyperplasia who developed maculopapular rash and rash on the extremities after 2 months of drug administration[66]. According to the FAERS database, SOCs not mentioned in the specification included endocrine system diseases, renal and urinary system diseases, various surgical and medical operations, and various congenital familial hereditary diseases. It should be noted that we conducted a revalidation of the results of the specification from a pharmacovigilance perspective for reproductive and breast disorders, psychiatric categories, and various neurological disorders. There is emerging evidence that finasteride both treats suppurative sweats in women[67] and reduces the incidence of bladder cancer[68,69]and induces type 2 diabetes[70], down-regulates androgen receptor expression in the renal cortex[71], forms blood clots[72], and alters intestinal microbial abundance[73,73,74] among other things. With regard to ‘endocrine disorders’ and ‘renal and urinary disorders’ it is noteworthy that almost all clinical trials did not include primary and secondary endpoint sensitisers for type 2 diabetes mellitus, renal dysfunction indicators. These unintended AEs raise concerns about the effects of finasteride on endocrine and renal disorders and the potential risk of severe endocrine disorders[75]. Further studies are necessary to investigate the prevalence of hormonal and metabolic changes and to evaluate the level of risk for individuals taking finasteride.

With respect to the timing of adverse events and final outcome, the probability of adverse events was greatest on days 0-31 of finasteride use in our analysis. Therefore, it is important to closely monitor potential risks associated with medication use over a one-month period. In addition, disability and life-threatening events such as reproductive tract hypoplasia, male and catastrophic reactions were also reported in our study, suggesting that comprehensive monitoring and management of AEs receiving finasteride medication should not be underestimated, especially in the treatment of male patients with androgenetic alopecia and prostatic hyperplasia[76].

From the perspective of gender subgroups, male patients reported a higher incidence of adverse events with finasteride compared to female patients. This gender difference can be understood from both sociological and biological perspectives. From the sociological point of view, firstly, the prevalence of AGA is on the rise globally, with 45.72% of men and 5.05% of women[77]. Furthermore, premenopausal women suffering from hair loss are mainly treated with minoxidil, oral contraceptives, and cosmetic products[78]. Finasteride, on the other hand, has been used to promote hair growth mainly after menopause[79,80]. Therefore, the use of finasteride has been found to be more likely to cause adverse events in male patients than in female patients. As a result, the population of women who use finasteride is relatively small. Secondly, women prefer hair transplants and cosmetics to increase scalp hair fullness[81]. Finally, the safety of oral finasteride in patients with a history of gynaecological malignancy has not yet been clarified, which makes the use of this drug in the treatment of androgenetic alopecia more limited, especially when compared to male patients[82]. Biologically, AGA, previously thought to have an autosomal dominant mode of inheritance, has a reduced epistasis in females[83]. This makes males more susceptible to AGA than females[84]. Furthermore, female follicle levels of type I and type II 5α-reductase and androgen receptors are half those of males. The percentage of females with a clinical diagnosis of AGA is then lower than that of males. In summary, we hypothesize that men experience more adverse events than women because of the aforementioned biological and sociological factors, and further investigation into the underlying mechanisms and reasons is warranted. At the age subgroup level, our study found that patients between the ages of 18 and 65 years were more likely to report adverse events involving finasteride than patients younger than 18 years and those older than or equal to 65 years. Because of the high correlation between AGA severity and age factors[85], AGA not only affects social functioning and emotional well-being in young men[86], but also affects sexual functioning and reduces quality of life in women aged 18-65 years[87]. Therefore, clinicians can kindly remind patients aged 18-65 years of the medication precautions when using it in the clinical setting, so that finasteride AEs will not go unnoticed in the non-medical setting.

## Limitation

It is crucial to recognize some constraints of our research, primarily arising from the fundamental features of pharmacovigilance databases. Firstly, up to 53.67% of the reports in our study originated from consumers (n=6203), given the potential for inaccurate or misleading content in reports submitted by non-medical professionals due to limitations in their medical knowledge. Second, our data analyses failed to cover a wide range of unquantified confounding variables that may affect adverse events (AEs), such as potential drug interactions, changes in treatment regimens, and results of laboratory tests and instrumental examinations. Finally, additional prospective studies are required to validate and clarify the relationship between finasteride and these adverse events. These studies should address issues such as data reporting inconsistencies, incomplete data, population heterogeneity, and other confounding factors that may impact the findings of the data analyses.

## Conclusions

We conducted pharmacovigilance analyses using actual data from the FAERS database, and the adverse reactions identified in our investigation largely aligned with those specified. Additionally, we detected adverse events not listed in the specifications, including post 5-alpha-reductase inhibitor syndrome, Peyronie’s disease, testicular microlithiasis, and catastrophic reactions. The identification of these strongly indicating AEs supplements the inherent limitation of the relatively small sample size in clinical studies of this drug. The results of this study help to inform the safe and rational use of the drug in the clinic.

## Data Availability

All relevant data are within the manuscript and its Supporting Information files.

## Acknowledgments

We would like to thank the participants and researchers of the FAERS database. We also acknowledge Medex UIMA 1.8.3 system for providing their platforms and contributors for uploading meaningful data.

## Author contributions

XZ and YY designed the study and edited the manuscript. SW performed the statistical analysis and drafted the manuscript. JC and ZW generated the figures for the manuscript. YL reviewed and edited the manuscript. All authors contributed to the article and approved the submitted version.

## Supporting information

**S1 Table. Four grid table.**

**S2 Table. Four major algorithms used for signal detection.**

**S3 Table. Signal strength of reports of finasteride administration at the SOC level**

**S4 Table. Signal strength of reports of finasteride administration in female at the PT level.**

**S5 Table. Signal strength of reports of finasteride administration in male at the PT level.**

**S6 Table. Signal strength of reports of finasteride administration at the PT level (age<18).**

**S7 Table. Signal strength of reports of finasteride administration at the PT level (age=18~65).**

**S8 Table. Signal strength of reports of finasteride administration at the PT level (age >=65).**

